# Neighborhood Deprivation and Racial Disparities in Metastatic Prostate Cancer at Diagnosis: A Population-Based Study in Ohio

**DOI:** 10.64898/2026.06.02.26354723

**Authors:** Julia Y. Payne, Stephen P. Rhodes, Jonathan Shoag, Michael B. Rothberg, Phuc Le, Jennifer Cullen, Holly Hartman

## Abstract

**Background:** Prostate cancer survival varies by stage at diagnosis, and Black men experience a disproportionate burden of advanced disease. We examined whether neighborhood deprivation, measured by Area Deprivation Index (ADI), contributes to racial differences in metastatic presentation.

**Methods:** We conducted a population-based study of men diagnosed with prostate cancer in the Ohio Cancer Incidence Surveillance System from 1996 to 2016. The primary endpoint was distant-stage disease at diagnosis. Generalized additive models assessed nonlinear associations of ADI and diagnosis year with metastatic risk. Inverse probability of treatment weighting (IPTW) models estimated odds ratios comparing Black with White men after sequential adjustment for diagnosis year, age, insurance, and ADI.

**Results:** Among 135,095 men, 18,690 were Black and 116,405 were White. Distant-stage disease occurred in 7.0% of Black men and 5.0% of White men. Black men had higher median ADI (60.9 vs. 47.3). Medicaid-insured men had the highest unadjusted odds of metastatic presentation (OR, 4.68; 95% CI, 4.13–5.31), exceeding uninsured men (OR, 2.91; 95% CI, 2.54–3.34). In IPTW models without age adjustment, the odds ratio decreased from 1.54 to 1.24 after adding insurance and ADI. In age-adjusted IPTW models, the odds ratio decreased from 1.79 to 1.41 after adding insurance and ADI. Generalized additive models showed increasing metastatic risk at higher ADI values and after 2008.

**Conclusions:** Neighborhood deprivation and insurance-related access explained part, but not all, of the excess odds of metastatic diagnosis among Black men.

**Impact:** Integrating ADI into cancer surveillance may improve identification of populations at risk for late-stage diagnosis.

## INTRODUCTION

Prostate cancer (PCa) remains one of the most common cancers among men in the United States, and survival varies substantially by stage at diagnosis. Population-based estimates show excellent survival for localized disease, whereas metastatic PCa at initial presentation is associated with substantially poorer survival (1–3). Because stage at diagnosis is a strong determinant of prognosis, racial disparities in advanced-stage presentation are a major public health concern, especially because Black men experience higher PCa incidence, mortality, and likelihood of advanced-stage disease than White men (2). Although ancestry-associated germline and somatic differences may contribute to variation in PCa risk and tumor biology, these factors do not fully explain population-level racial disparities in stage at diagnosis or outcomes (4,5). Because race reflects social and structural exposures in addition to ancestry, racial differences in metastatic presentation should be interpreted in the context of healthcare access, screening and diagnostic pathways, and broader social conditions (2,6–11).

Social and economic conditions have long been recognized as contributors to cancer disparities, although recent literature increasingly frames these factors as social determinants of health (SDOH). SDOH are the nonmedical conditions that shape health, including economic stability, education, healthcare access, neighborhood environment, and social context (12). In oncology, these factors are associated with differences in early detection, stage at diagnosis, and survival outcomes (13). National cancer-disparities frameworks also emphasize structural and neighborhood-level factors as key drivers of unequal cancer burden (14,15). In PCa, social and economic disadvantage may contribute to metastatic presentation through barriers to primary care, insurance coverage, and timely diagnostic follow-up. Systematic reviews and population-based studies have linked SDOH and neighborhood deprivation to differences in stage at diagnosis, treatment, mortality, and survival among Black and White patients with PCa (13–16). These findings suggest that neighborhood deprivation may affect whether PCa is detected before metastatic progression (13,15).

The Area Deprivation Index (ADI) provides a standardized, geographically linked marker of neighborhood-level socioeconomic disadvantage (16,17). ADI is useful for registry-based cancer disparities research because cancer registries often lack detailed individual-level measures of income, wealth, education, housing stability, transportation, and other SDOH. Unlike single measures of socioeconomic status, ADI summarizes multiple neighborhood characteristics, including income, education, employment, and housing quality, at the census block group level (16,17). Census block groups offer greater geographic precision than census tracts or ZIP codes, which may contain more heterogeneous populations and obscure local variation in deprivation. This finer geographic resolution may better approximate the social and economic context surrounding a patient’s residence at diagnosis. Prior studies have linked neighborhood deprivation with PCa risk, mortality, and survival, including evidence that deprivation may contribute to racial differences in PCa outcomes (18–23). Equal-access healthcare studies have shown attenuation of racial differences in PCa outcomes, supporting the importance of access and structural context in interpreting race-based disparities (24,25). Therefore, ADI was selected as the primary SDOH measure for this study because it captures broader neighborhood disadvantage and can be linked to registry data at a granular geographic level.

Using the Ohio Cancer Incidence Surveillance System (OCISS), a statewide cancer registry administered by the Ohio Department of Health, we examined whether neighborhood socioeconomic deprivation, measured by ADI, was associated with metastatic PCa at initial diagnosis among Black and White men in Ohio from 1996 to 2016. We hypothesized that higher ADI values would be associated with greater odds of metastatic presentation and that accounting for ADI would reduce the elevated odds of distant-stage PCa among Black men compared with White men.

## METHODS

We conducted a population-based retrospective cohort study using data from OCISS, a statewide cancer registry that collects cancer incidence data reported by clinics, hospitals, pathology laboratories, and other healthcare facilities throughout Ohio. Registry records include diagnosis date, stage at diagnosis, anatomic site, tumor grade, patient demographics, and health insurance status. Access to OCISS data was approved by the Ohio Department of Health Institutional Review Board, as required for use of registry data, and informed consent was not required for using de-identified data.

The study population included men aged 18 years or older with primary PCa diagnosis between January 1, 1996, and December 31, 2016. The 1996–2016 study period represented the approved OCISS analytic window with required registry variables for cohort identification, stage classification, insurance status, race, and ADI linkage. Exclusion criteria included missing ADI values, unstaged disease at diagnosis, age younger than 18 years, and race categories other than Black or White, including multiple, other, or unknown race. The final analytic cohort included 135,095 men with PCa and available ADI data. Subject demographics included age at diagnosis, sex, race, ethnicity, insurance status, diagnosis year, cancer stage at diagnosis, and ADI value. All included subjects were male, as the cohort was restricted to men diagnosed with PCa. Body weight was not captured in OCISS and was not analyzed.

The primary outcome was distant-stage PCa at initial diagnosis, used as a marker of metastatic presentation. Men with localized, regional, or in situ disease were categorized as having nonmetastatic disease. Race was categorized as Black or White based on registry data. Covariates included age at diagnosis, diagnosis year, insurance status, and ADI. Insurance status was categorized as private insurance, Medicaid/public health insurance, Medicare, Military/Veterans Affairs, not insured, or unknown.

Neighborhood socioeconomic deprivation was measured using ADI, a census-block group level index based on socioeconomic indicators including income, poverty, education, employment, and housing conditions. Higher ADI values indicate greater neighborhood deprivation. ADI values were assigned based on patient’s residential address at diagnosis and obtained from the Neighborhood Atlas using the sociome package in R. ADI was analyzed as a continuous variable in regression models and grouped into deciles in descriptive analyses.

Descriptive statistics were used to characterize the cohort overall and by race and metastatic status at diagnosis. Temporal trends in the proportion of men diagnosed with distant-stage PCa were examined across diagnosis years. Generalized additive models (GAMs) were used to evaluate potential nonlinear associations between ADI, diagnosis year, and the probability of distant-stage PCa at diagnosis. Models were adjusted for race, age at diagnosis, insurance status, diagnosis year, and ADI. Estimated smooth functions were visualized to assess the shape and magnitude of associations across the range of ADI values and diagnosis years. Stratified analyses evaluated whether associations varied across diagnosis periods and by insurance status.

To address measured covariate imbalance between Black and White men, inverse probability of treatment weighting (IPTW) was applied using propensity scores estimated from logistic regression, with race as the exposure variable. Three sequential propensity score models were constructed to evaluate whether balancing on measured covariates reduced the higher odds of distant-stage diagnosis among Black men compared with White men. Model A included diagnosis year and age at diagnosis; Model B added insurance status; and Model C added ADI. Covariate balance before and after weighting was assessed using standardized mean differences, with values less than 0.1. Weighted logistic regression models were then used to estimate odds ratios for distant-stage PCa at diagnosis among Black men compared with White men.

This study used existing registry data, and subjects were not randomly assigned to exposure groups. Group assignment was based on race as recorded in OCISS and neighborhood deprivation as measured by ADI. Blinding was not performed because the primary outcome, distant-stage PCa at diagnosis, was obtained from registry-recorded stage data before statistical analysis. No formal power calculation was performed because the sample size was determined by all eligible PCa cases in OCISS during the approved study period.

All statistical analyses were performed using R version 3.4.2. Data management, statistical modeling, and visualization were conducted using reproducible R Markdown workflows with version-controlled scripts documenting analytic decisions and model development. GAMs were fit using the mgcv package, propensity score models and contrasts were estimated using the marginaleffects package, and data management and visualization used the tidyverse package suite. Analytic code is available from the corresponding author upon reasonable request.

## Data Availability Statement

The Ohio Cancer Incidence Surveillance System data used in this study are maintained by the Ohio Department of Health and are available to qualified researchers through a formal data use agreement (https://odh.ohio.gov/know-our-programs/ohio-cancer-incidence-surveillance-system). Area Deprivation Index data are publicly available from the Neighborhood Atlas (https://www.neighborhoodatlas.medicine.wisc.edu/) and were accessed programmatically using the sociome R package (https://CRAN.R-project.org/package=sociome). Analytic code is available from the corresponding author upon reasonable request.

## RESULTS

Our study included 135,095 men diagnosed with PCa in Ohio from 1996 to 2016, of which 18,690 (13.8%) were Black and 116,405 (86.2%) were White (Table 1). The median age of diagnosis for the cohort was 67 years [IQR: 61–73], with Black men being diagnosed at a younger median age (65 years, IQR: 58–71) compared to White men (67 years, IQR: 61–74). When comparing insurance coverage in Table 1, Black men had a lower proportion of Medicare coverage (41%) compared to White men (48%), and a higher proportion of Medicaid/Public Health insurance (6.5% vs. 1.3% for White men).

**Table 1:** Summary of characteristics by race.

| Variables | OCISS Cohort |  |  |
| --- | --- | --- | --- |
|  | Total | White | Black |
| **Sample Size** | 135095 (100%) | 116405 (86.2%) | 18690 (13.8%) |
| **Race/Ethnicity** |  |  |  |
| **Non-Hispanic** | 122715 (90.8%) | 105641 (91%) | 17074 (91%) |
| **Hispanic** | 684 (0.5%) | 639 (0.6%) | 45 (0.2%) |
| **Diagnosis Year,**<br>**median (IQR)** | 2006<br>(2001-2011) | 2006<br>(2001-2011) | 2007<br>(2002-2012) |
| **Diagnosis Age,**<br>**median (IQR)** | 67<br>(61-73) | 67<br>(61-74) | 65<br>(58-71) |
| **Insurance** |  |  |  |
| **Private Insurance** | 47950 (35.5%) | 41157 (35%) | 6793 (36%) |
| **Medicaid/**<br>**Public Health** | 2732 (2%) | 1522 (1.3%) | 1210 (6.5%) |
| **Medicare** | 63102 (46.7%) | 55439 (48%) | 7663 (41%) |
| **Military/VA** | 2435 (1.8%) | 1701 (1.5%) | 734 (3.9%) |
| **Not Insured** | 3324 (2.5%) | 2712 (2.3%) | 612 (3.3%) |
| **OH Stage <sup>a</sup>** |  |  |  |
| **Distant** | 6598 (4.9%) | 5310 (5%) | 1288 (7%) |
| **In situ** | 54 (0.04%) | 45 (0.04%) | 9 (0.05%) |
| **Localized** | 114346 (84.6%) | 98803 (85%) | 15543 (83%) |
| **Regional** | 14097 (10.4%) | 12247 (11%) | 1850 (10%) |
| **Area Deprivation Index,**<br>**median (IQR)** | 48.3<br>(42.0-55.8) | 47.3<br>(41.4-53.5) | 60.9<br>(51.5-69.1) |

<sup>a</sup> OH Stage (Ohio Field): single stage field that can be used across all years that takes into account reported stage data and year of diagnosis.

Among the participants, 6,598 (4.9%) were diagnosed with distant metastatic PCa at initial diagnosis, while 114,346 (84.6%) were diagnosed with localized PCa, and 14,097 (10.4%) had regional PCa (Table 1). The proportion of distant-stage disease was higher among Black men than White men (7.0% vs. 5.0%). Additionally, Black men had higher socioeconomic deprivation, as reflected by the median ADI score: 60.9 [IQR: 51.5–69.1] for Black men versus 47.3 (IQR: 41.4–53.5) for White men (Table 1). The proportion of men diagnosed with distant-stage disease increased across ADI deciles, and Black men had a higher proportion than White men at nearly every ADI decile (Figure 1).

**Figure 1:**
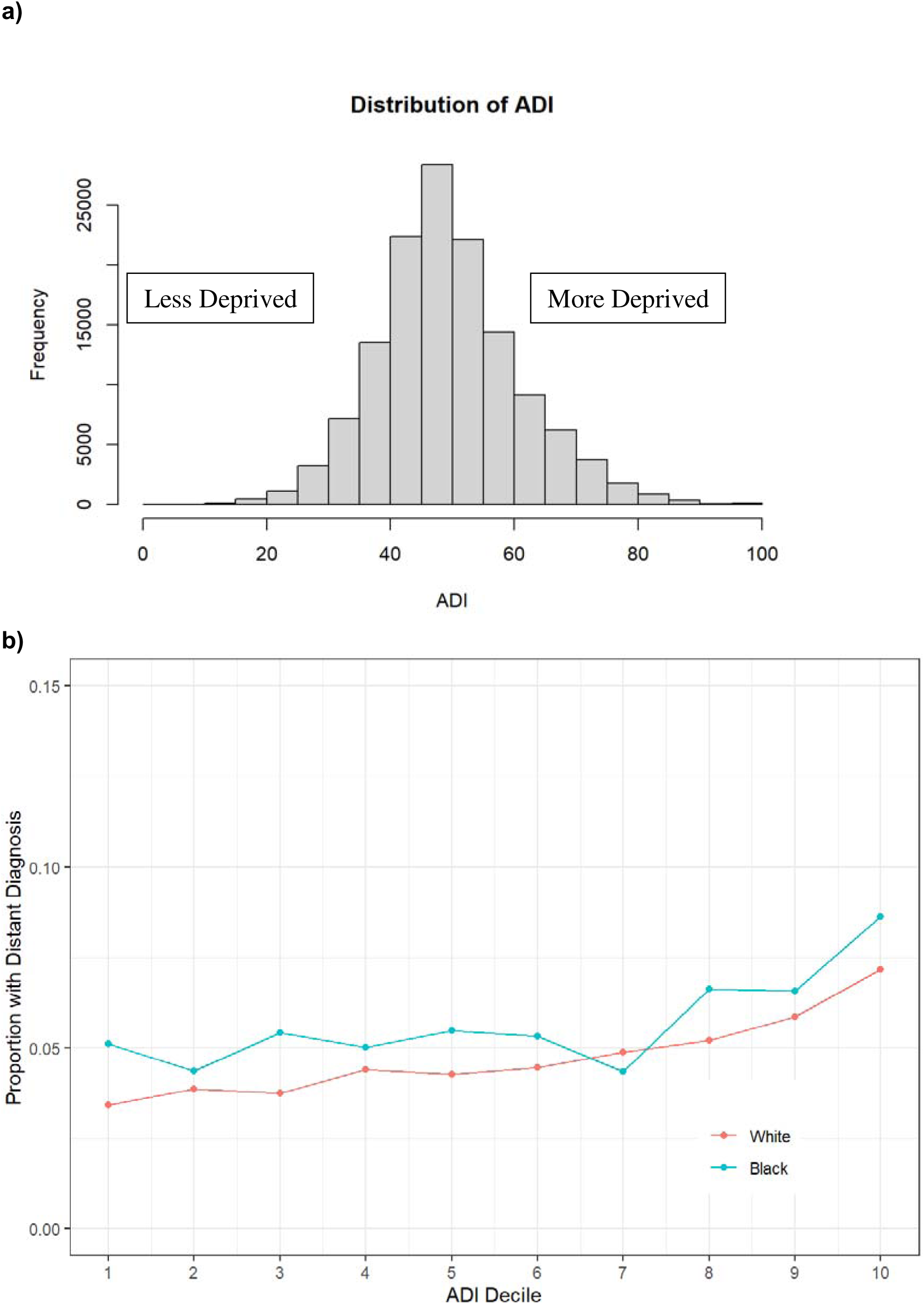
a) Distribution of Area Deprivation Index in the OCISS cohort and b) distribution of Area Deprivation Index decile separated by race.

Consistent with these descriptive patterns, GAMs showed that the predicted probability of distant-stage PCa at diagnosis was relatively stable at lower ADI values and increased more steeply at mid-to-high levels of neighborhood deprivation (Figure 2). Men with an ADI of 80 had approximately twice the predicted probability of distant-stage disease compared with men with an ADI of 40.

**Figure 2:**
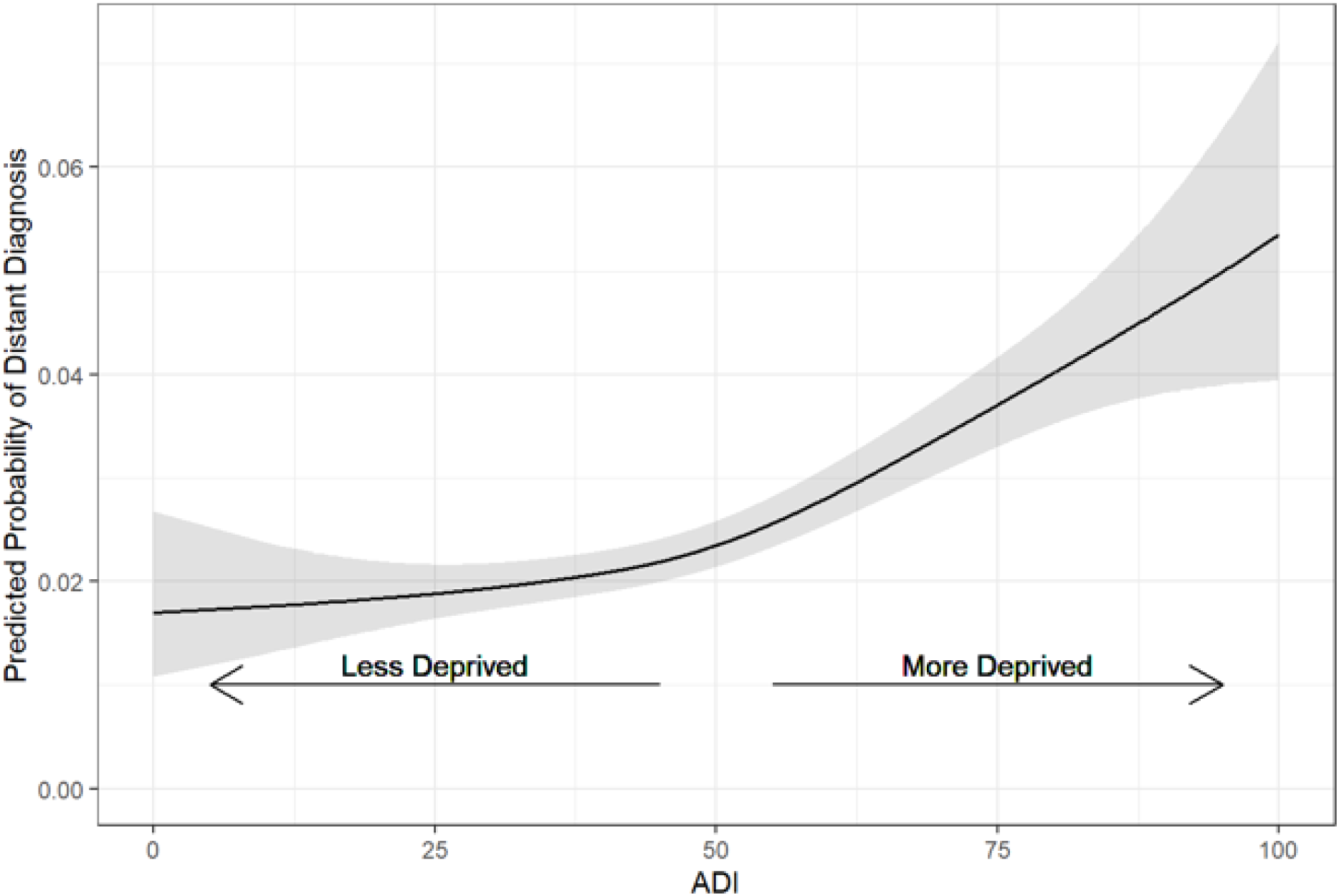
Predicted risk of distant diagnosis of prostate cancer based on Area Deprivation Index

The temporal patterns also differed by race. Across the study period, both Black and White men showed declining predicted probabilities of distant-stage PCa through the early 2000s, followed by an increase beginning around 2008. The later increase appeared steeper among White men. However, Black men maintained a higher absolute predicted probability of distant-stage disease throughout the study period (Suppl. Figures 2 & 3). Black men also had higher predicted risk of distant-stage PCa at earlier ages compared with White men (Suppl. Figure 3).

In unadjusted analyses, Black men had higher odds of distant-stage PCa at diagnosis compared with White men (OR, 1.55; 95% CI, 1.45–1.65). Insurance status was also strongly associated with distant metastatic PCa at diagnosis (Table 2). Compared to men with private insurance, those insured through Medicaid or other public health programs exhibited more than fourfold higher odds of presenting with metastatic disease (OR 4.68, 95% CI 4.13–5.31). Elevated odds of metastatic presentation were also observed among uninsured men (OR 2.91, 95% CI 2.54–3.34), those insured through Medicare (OR 2.21, 95% CI 2.08–2.35), and those receiving care through the Military or Veterans Affairs health systems (OR 1.64, 95% CI 1.35–2.0). Men with unknown insurance status also demonstrated increased odds of metastatic disease at diagnosis (OR 1.41, 95% CI 1.29–1.56).

**Table 2.** Unadjusted odds ratios for distant-stage prostate cancer at initial diagnosis by race and insurance status.

| Predictor | Reference group | OR | 95% CI |
| --- | --- | --- | --- |
| **Race** |  |  |  |
| Black | White | 1.55 | 1.45–1.65 |
| **Insurance status** |  |  |  |
| Medicaid/Public Health | Private insurance | 4.68 | 4.13–5.31 |
| Medicare | Private insurance | 2.21 | 2.08–2.35 |
| Military/VA | Private insurance | 1.64 | 1.35–2.00 |
| Not insured | Private insurance | 2.91 | 2.54–3.34 |
| Unknown insurance | Private insurance | 1.41 | 1.29–1.56 |

*OR = odds ratio; CI = confidence interval; VA = Veterans Affairs. Odds ratios are unadjusted.*

To further evaluate whether measured covariates accounted for higher odds of distant-stage diagnosis among Black men, we used IPTW propensity score models and balanced groups on factors such as age, year of diagnosis, insurance type, and ADI (Table 3). Before weighting, ADI was the most imbalanced measured covariate between Black and White men (standardized mean difference [SMD], 1.084). After IPTW, ADI balance improved substantially (SMD, 0.034) (Table 3). We included additional models to account for differences in age at diagnosis since younger age at diagnosis may be associated with more aggressive disease presentation. Given the younger median age at diagnosis among Black men compared with White men, sequential IPTW models were estimated with and without age adjustment to assess whether age at diagnosis influenced the difference in odds of distant-stage PCa between Black and White men.

**Table 3:** Balancing of variables in propensity score models using inverse probability of treatment weighting (IPTW).

| **Propensity Score Models** |  |  |
| --- | --- | --- |
| **A)** | **Differences Unadjusted** | **Differences Adjusted** |
| **Diagnosis Year** | -0.284 | 0.011 |
| **Diagnosis Age** | 0.102 | -0.009 |
| **B)** |  |  |
| **Diagnosis Year** | -0.284 | 0.020 |
| **Diagnosis Age** | 0.102 | -0.010 |
| **Insurance** |  |  |
| **Private Insurance** | 0.010 | 0.007 |
| **Medicaid/Public Health** | 0.052 | 0.000 |
| **Medicare** | -0.066 | -0.007 |
| **Military/VA** | 0.025 | 0.000 |
| **Not Insured** | 0.009 | -0.002 |
| **C)** |  |  |
| **Diagnosis Year** | -0.284 | -0.032 |
| **Diagnosis Age** | 0.102 | 0.026 |
| **Insurance** |  |  |
| **Private Insurance** | 0.010 | 0.016 |
| **Medicaid/Public Health** | 0.052 | 0.001 |
| **Medicare** | -0.066 | -0.020 |
| **Military/VA** | 0.025 | 0.001 |
| **Not Insured** | 0.009 | 0.000 |
| **ADI** | 1.084 | 0.034 |

In models without age adjustment, the odds ratio comparing Black men with White men decreased from 1.54 (95% CI, 1.45–1.64) after balancing for diagnosis year to 1.24 (95% CI, 1.13–1.35) after adding insurance and ADI. In age-adjusted models, the corresponding odds ratios were higher, decreasing from 1.79 (95% CI, 1.68–1.91) after balancing for diagnosis year and age to 1.41 (95% CI, 1.29–1.55) after adding insurance and ADI. Across both modeling approaches, accounting for insurance status and ADI reduced the magnitude of elevated odds among Black men by approximately 56% in models without age adjustment and by 48% in age-adjusted models, although higher odds of distant-stage diagnosis persisted (Figure 3).

**Figure 3:**
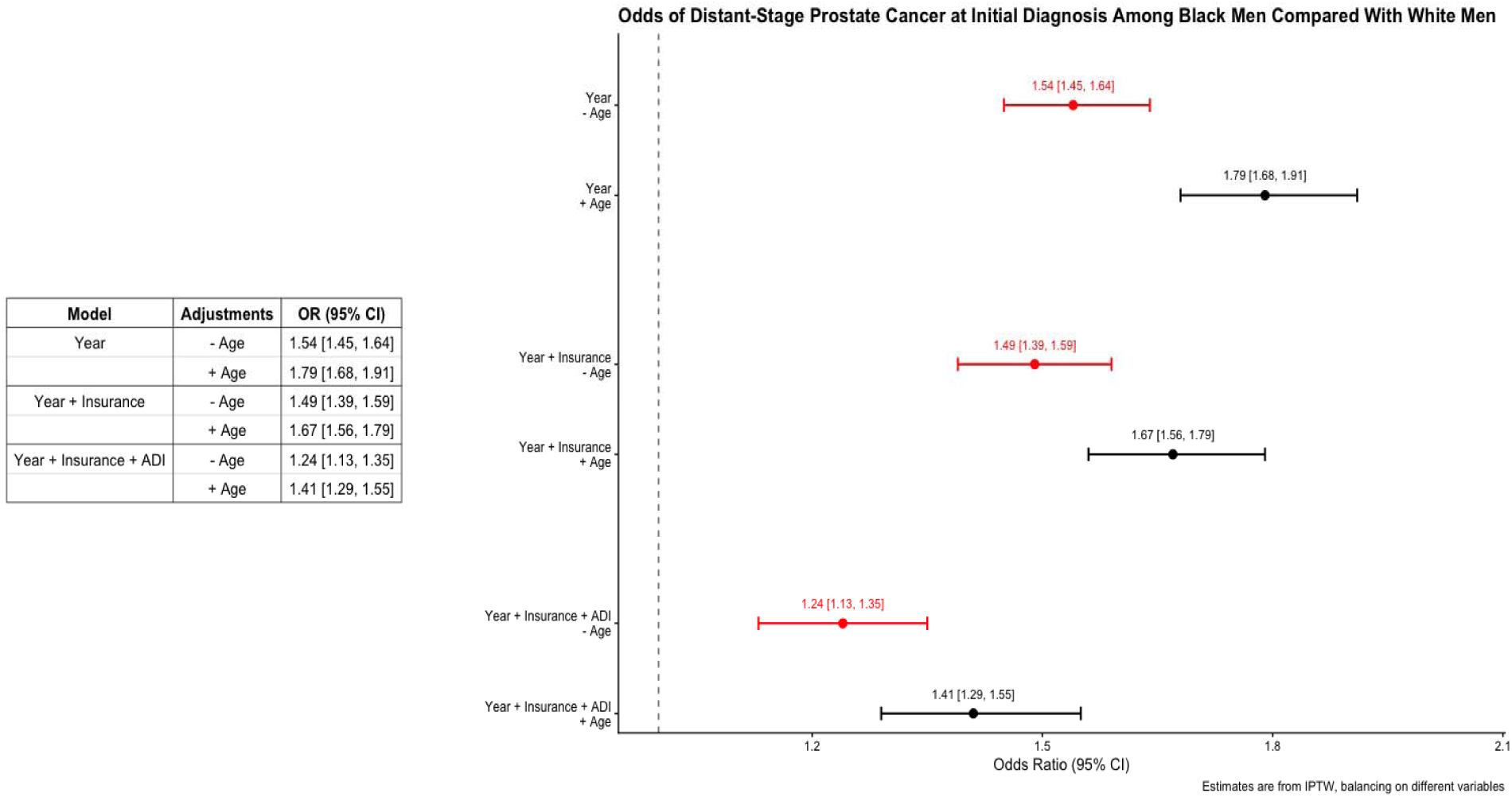
Propensity Score Models: Forest plot using IPWT showing odds ratio of distant PCa during initial diagnosis for Black and White men. The first propensity score model included only balanced for diagnosis year & age. The second model also balanced on insurance type. The third model also balanced on ADI.

## DISCUSSION

Using a statewide, population-based cancer registry linked to census block group level ADI, this study identified several findings relevant to PCa disparities and cancer surveillance. Among 135,095 Ohio men diagnosed with PCa between 1996 and 2016, Black men had higher odds of distant-stage disease at initial diagnosis than White men, and the probability of metastatic presentation increased with greater neighborhood deprivation. The main contribution of this study is that ADI was not only associated with metastatic presentation but also accounted for a meaningful portion of the elevated odds of distant-stage disease among Black men after balancing on measured covariates. These findings support interpreting racial differences in metastatic PCa presentation within the context of neighborhood socioeconomic disadvantage, insurance-related barriers, and unequal opportunities for early detection rather than race alone as a biological explanation.

Neighborhood deprivation was central to the observed difference in metastatic presentation. Before weighting, ADI was the most imbalanced measured covariate between Black and White men, with a standardized mean difference of 1.084, indicating substantially greater neighborhood deprivation among Black men. After IPTW including ADI, covariate balance improved substantially (standardized mean difference, 0.034). In the propensity score model balanced for diagnosis year and age, Black men had 79% higher odds of distant-stage PCa at diagnosis compared with White men (OR, 1.79; 95% CI, 1.68-1.91). Adding insurance status reduced the odds ratio to 1.67 (95% CI, 1.56-1.79), and further adding ADI reduced it to 1.41 (95% CI, 1.29-1.55). Together, insurance status and ADI reduced the magnitude of elevated odds among Black men by approximately one-half, suggesting that measured neighborhood and access-related factors explain an important component of metastatic-stage differences, although the persistence of elevated odds indicates that other unmeasured variables also remain important.

Temporal trends in our study suggest that metastatic presentation changed over the study period in ways that affected both racial groups. Metastatic PCa at initial diagnosis declined through the early 2000s but increased after approximately 2008 among both Black and White men. This timing overlaps with PSA screening recommendations that moved away from routine screening. This pattern is consistent with studies showing increasing metastatic PCa incidence after the 2008 and 2012 recommendations against routine PSA screening (26–30). However, we did not have individual-level PSA screening history, biopsy history, or diagnostic follow-up, and therefore our findings cannot establish that screening-policy changes caused the observed increase in metastatic presentation. These findings should instead be interpreted as observations suggesting that reduced screening intensity may have affected stage at diagnosis overall while pre-existing inequities likely contributed to persistently higher metastatic risk among Black men and men living in more deprived neighborhoods (31,32).

The association between insurance status and metastatic presentation should also be interpreted cautiously. In unadjusted analyses, men insured through Medicaid or public health programs had higher odds of metastatic presentation than uninsured men. This should not be interpreted as evidence that being uninsured is protective. Rather, insurance status recorded at diagnosis is an imperfect marker of longitudinal healthcare access and may reflect coverage obtained late in the diagnostic pathway (33–35). Medicaid coverage may identify patients with greater socioeconomic disadvantage, comorbidity, or enrollment triggered by cancer-related diagnosis (36,37). Medicaid coverage also does not guarantee timely access to PSA screening or biopsy because access may still be constrained by other healthcare barriers (34,36). The uninsured group, however, may include younger or healthier men, men with intermittent coverage, or men with limited access to diagnostic workup. Therefore, the higher risk observed among Medicaid-insured men may reflect greater socioeconomic and healthcare-access barriers rather than a harmful effect of Medicaid coverage itself. This interpretation is consistent with prior studies showing that lack of private insurance, including Medicaid or no insurance, is associated with later-stage disease and worse outcomes, and that Medicaid expansion or Medicaid coverage alone may not fully remove barriers to higher quality of care (33–37).

The persistence of elevated metastatic risk among Black men after adjustment for insurance status and ADI indicates that the measured social determinants available in OCISS captured only part of the difference in metastatic presentation. Residual differences may reflect unmeasured factors such as PSA testing, transportation barriers, trust in the healthcare system, and comorbidity burden. Although biological and genomic contributors cannot be evaluated in this registry-based analysis, the persistence of higher odds after adjustment should not be interpreted as evidence that racial differences are biologically determined. Rather, it underscores the need for more complete measurement of individual and neighborhood level determinants of PCa detection (24,25,31,38,39).

Several limitations should be considered. First, OCISS is a population-based registry and lacks detailed individual-level information on income, education, occupation, comorbidities, healthcare utilization, PSA screening history, or time from first abnormal test to diagnosis. Second, ADI captures neighborhood-level socioeconomic conditions and may not represent the individual socioeconomic circumstances of every resident within a census block group. Although ADI provides greater geographic granularity than ZIP code or county level measures, it remains subject to misclassification. In addition, ADI was assigned based on residential address at diagnosis using currently available census block group estimates, which may not fully align with neighborhood conditions during the entire study period from 1996-2016. Third, this analysis was limited to Black and White men because of the small numbers and missingness in other racial and ethnic categories, limiting generalizability. Fourth, propensity score weighting improved balance across measured covariates but cannot eliminate residual confounding from variables not captured in the registry. The reduction in odds after adding insurance status and ADI suggests that these measured social determinants explain part of the elevated odds among Black men, but the analysis cannot establish causality.

Despite these limitations, the findings have practical implications for cancer prevention and control. Efforts to reduce metastatic PCa should not focus only on individual screening decisions but should also address the neighborhoods and healthcare systems in which delayed detection occurs. Men living in high-ADI neighborhoods, Medicaid-insured men, and Black men may benefit from targeted outreach, patient navigation after abnormal PSA results, and policies that improve access to timely diagnostic evaluation. In conclusion, this statewide analysis demonstrates that neighborhood deprivation is associated with metastatic PCa at diagnosis and that incorporating ADI into propensity score models reduced, but did not eliminate, the elevated odds of distant-stage disease among Black men compared with White men. These results support integrating neighborhood-level deprivation measures into cancer surveillance and health-equity interventions aimed at reducing late-stage PCa presentation.

## SUPPLEMENTAL FIGURES

**Supplementary Figure 1.**
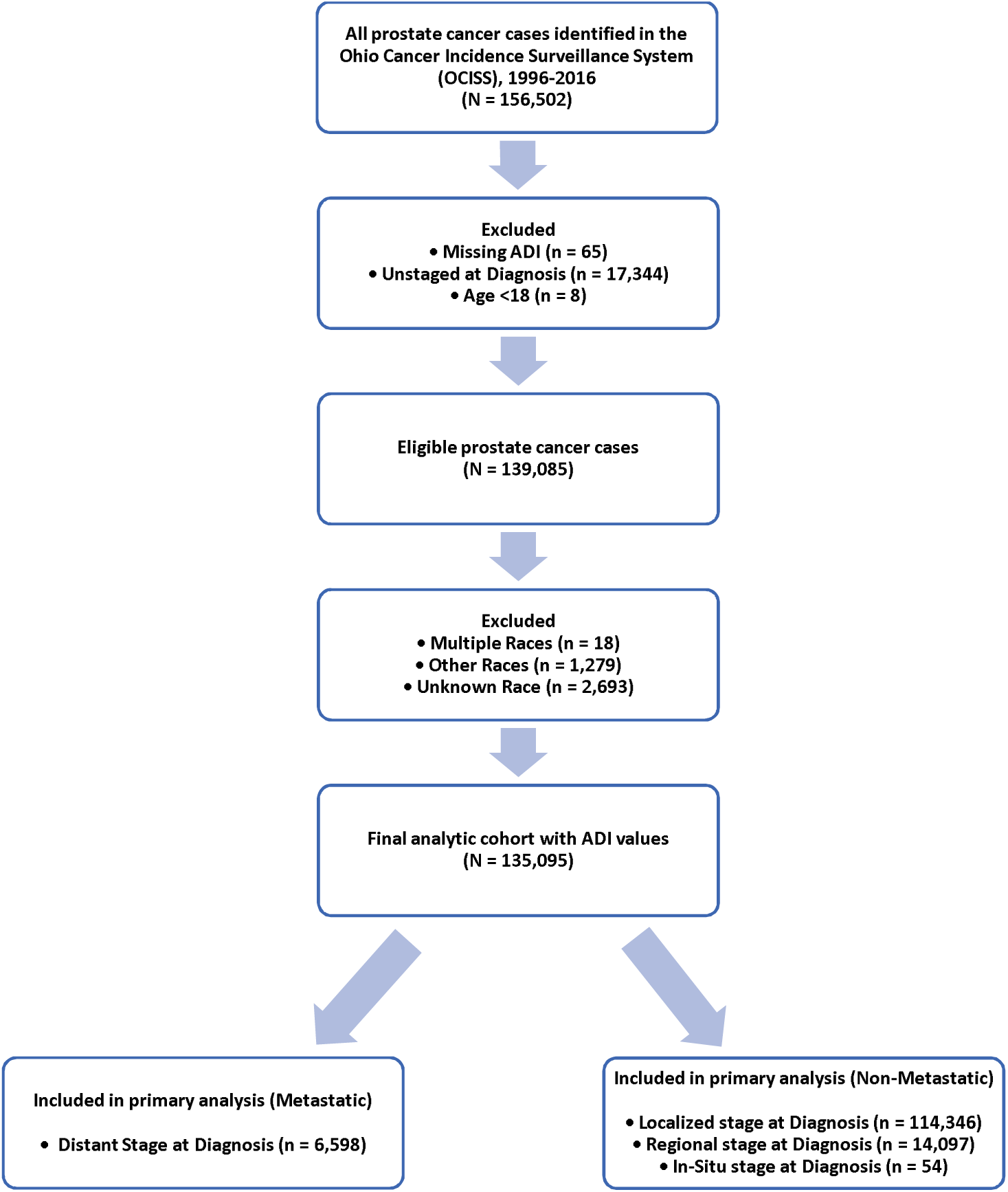
Flow Diagram of Study Cohort Selection

**Supplementary Figure 2:**
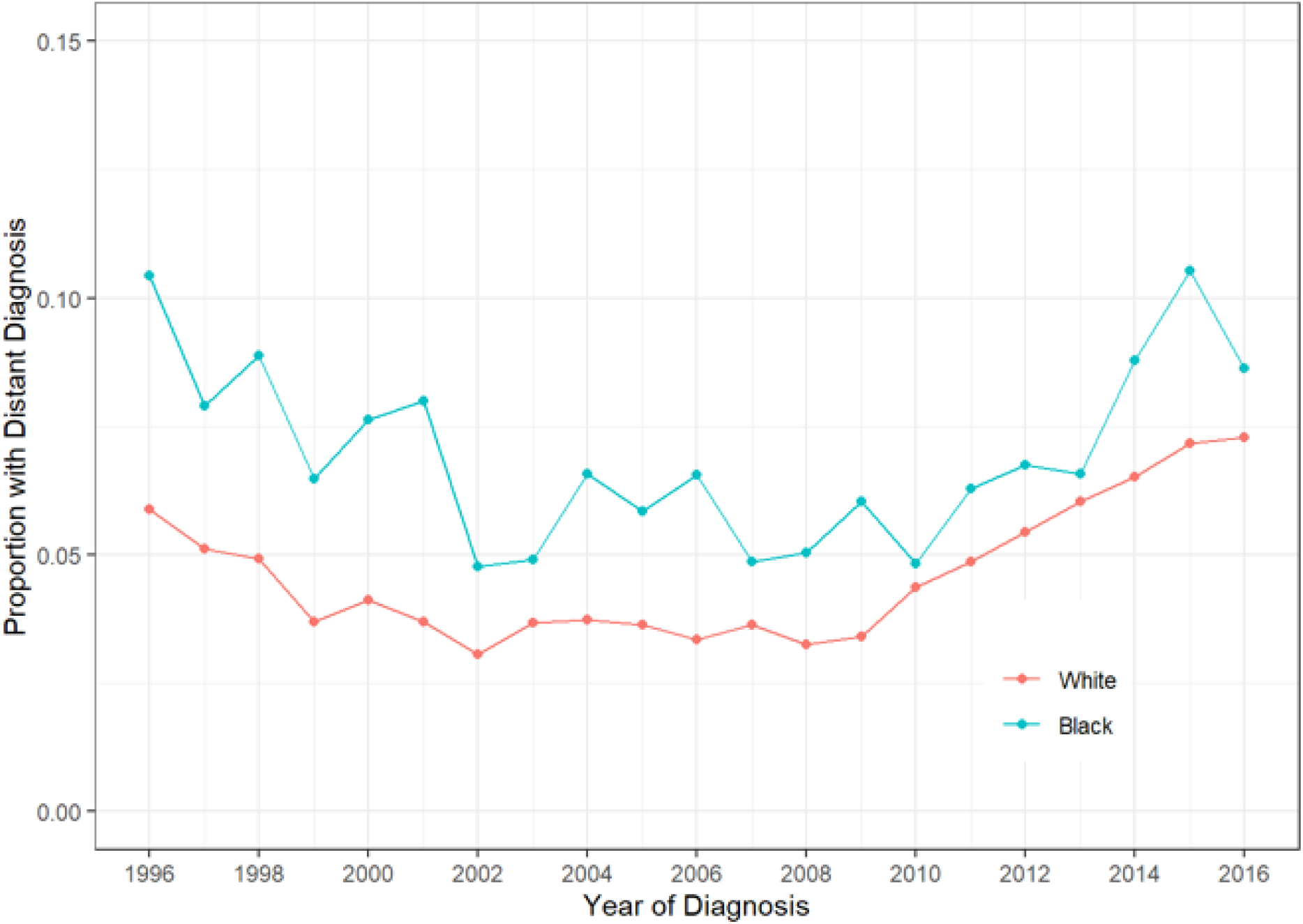
Distribution of initial distant diagnosis of prostate cancer by year of diagnosis separated by race.

**Supplementary Figure 3:**
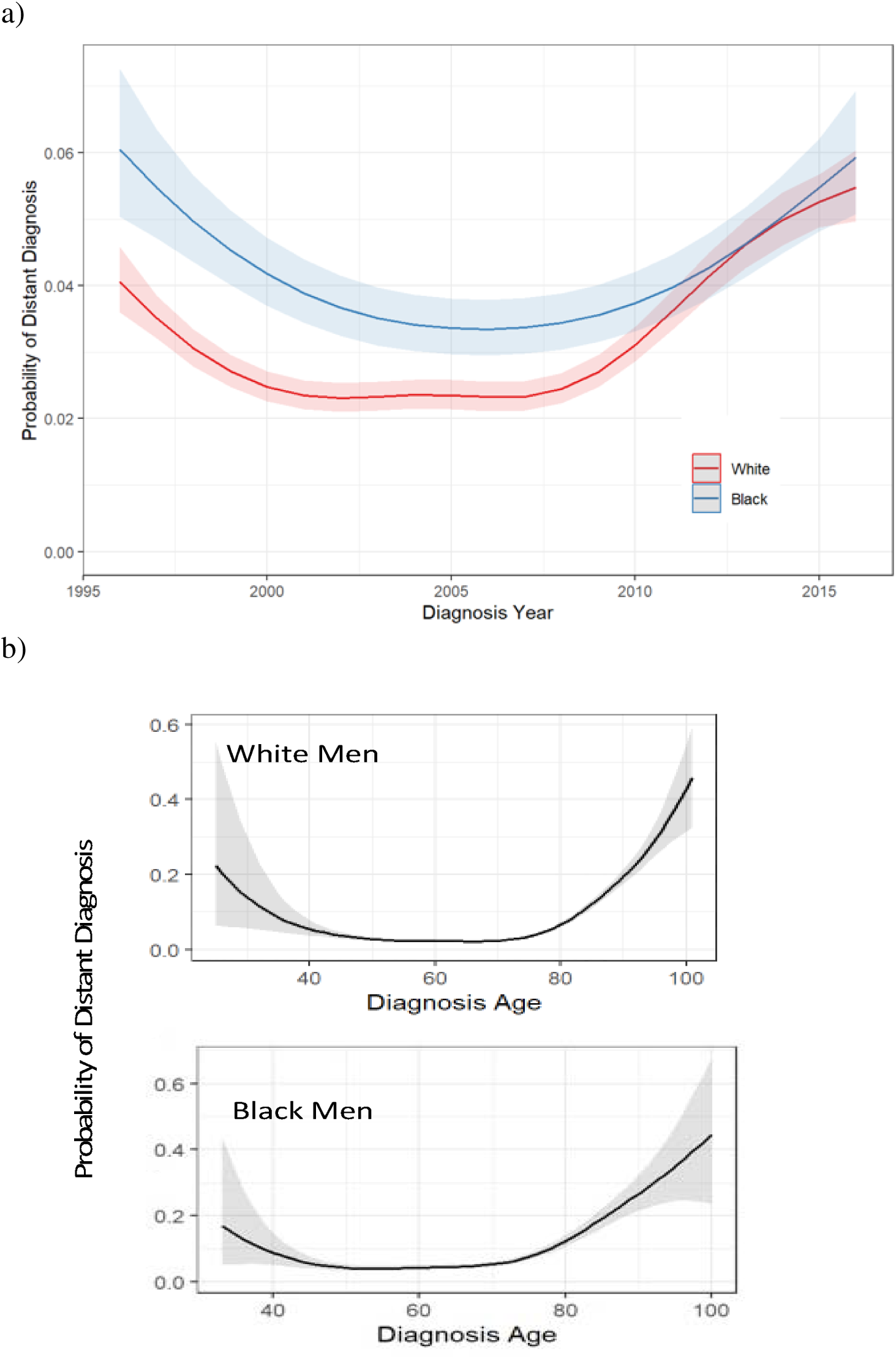
Trend of risk of distant diagnosis of prostate cancer by a) diagnosis year separated by race & b) diagnosis age separated by race.

## Notes

**Conflict of Interest:** The authors declare no potential conflicts of interest

### Competing Interest Statement

The authors have declared no competing interest.

### Author Declarations

Access to Ohio Cancer Incidence Surveillance System (OCISS) data was approved by the Ohio Department of Health Institutional Review Board, as required for use of registry data, and informed consent was not required for using de-identified data.

