## Supplementary Table 1 for "Neighborhood Deprivation and Racial Disparities in Metastatic Prostate Cancer at Diagnosis: A Population-Based Study in Ohio"

**Supplementary Table 1.** Covariate Balance Before and After Inverse Probability of Treatment Weighting Across Sequential Propensity Score Models, Ohio Cancer Incidence Surveillance System, 1996–2016

| **Propensity Score Model** | **Covariate** | **SMD Before Weighting** | **SMD After Weighting** |
| --- | --- | --- | --- |
| **Model A** |  |  |  |
|  | Diagnosis year | −0.284 | 0.011 |
|  | Diagnosis age | 0.102 | −0.009 |
| **Model B** |  |  |  |
|  | Diagnosis year | −0.284 | 0.020 |
|  | Diagnosis age | 0.102 | −0.010 |
|  | Private insurance | 0.010 | 0.007 |
|  | Medicaid/public health | 0.052 | 0.000 |
|  | Medicare | −0.066 | −0.007 |
|  | Military/VA | 0.025 | 0.000 |
|  | Not insured | 0.009 | −0.002 |
| **Model C** |  |  |  |
|  | Diagnosis year | −0.284 | −0.032 |
|  | Diagnosis age | 0.102 | 0.026 |
|  | Private insurance | 0.010 | 0.016 |
|  | Medicaid/public health | 0.052 | 0.001 |
|  | Medicare | −0.066 | −0.020 |
|  | Military/VA | 0.025 | 0.001 |
|  | Not insured | 0.009 | 0.000 |
|  | ADI | 1.084 | 0.034 |

Abbreviations: ADI, Area Deprivation Index; IPTW, inverse probability of treatment weighting; SMD, standardized mean difference; VA, Veterans Affairs.

Three sequential propensity score models were constructed with race (Black vs White) as the exposure variable. Model A balanced on diagnosis year and age at diagnosis. Model B added insurance status. Model C added Area Deprivation Index. SMD values <0.1 indicate adequate balance.
