## Supplementary Figures for "Neighborhood Deprivation and Racial Disparities in Metastatic Prostate Cancer at Diagnosis: A Population-Based Study in Ohio"

**SUPPLEMENTAL FIGURES**

**Supplementary Figure 1.** Flow Diagram of Study Cohort Selection

All prostate cancer cases identified in the Ohio Cancer Incidence Surveillance System (OCISS), 1996-2016 (N = 156,502)

Excluded • Missing ADI (n = 65) • Unstaged at Diagnosis (n = 17,344) • Age <18 (n = 8)

Eligible prostate cancer cases (N = 139,085)

Excluded • Multiple Races (n = 18) • Other Races (n = 1,279) • Unknown Race (n = 2,693)

Final analytic cohort with ADI values (N = 135,095)

Included in primary analysis (Metastatic) • Distant Stage at Diagnosis (n = 6,598)

Included in primary analysis (Non-Metastatic) • Localized stage at Diagnosis (n = 114,346) • Regional stage at Diagnosis (n = 14,097) • In-Situ stage at Diagnosis (n = 54)

**Supplementary Figure 2**: Distribution of initial distant diagnosis of prostate cancer by year of diagnosis separated by race.

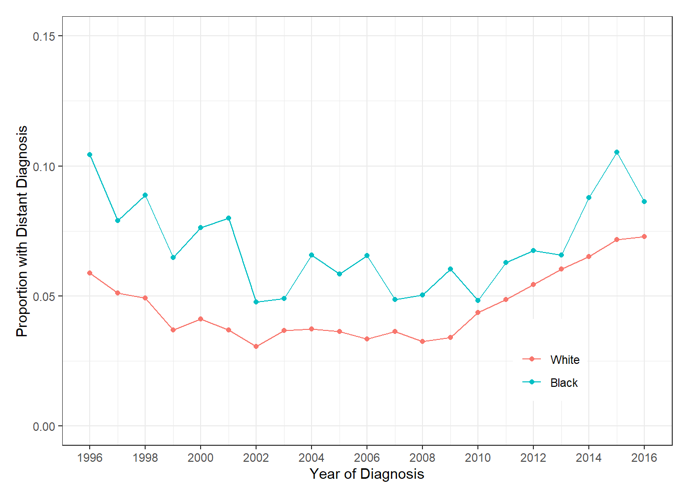

**Supplementary Figure 3**: Trend of risk of distant diagnosis of prostate cancer by a) diagnosis year separated by race & b) diagnosis age separated by race.

a)

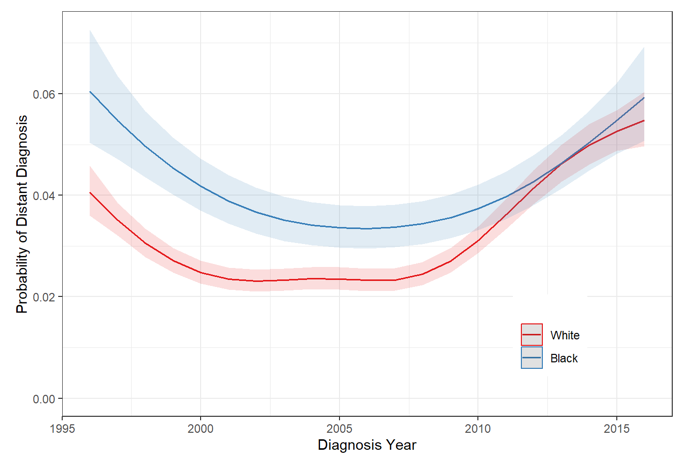

b)

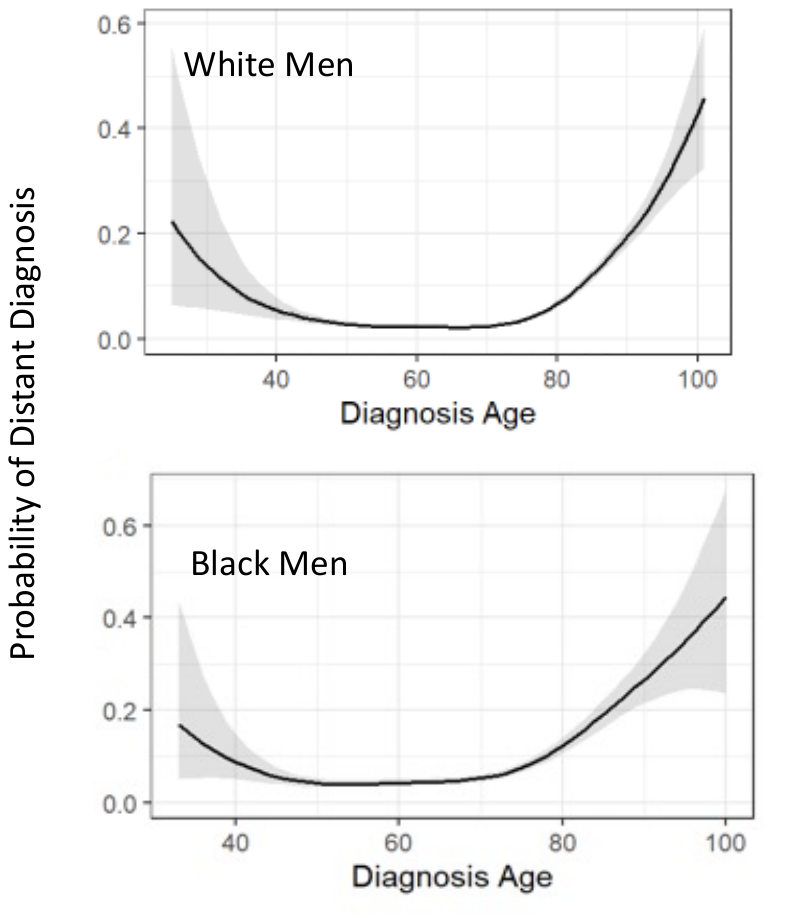
